# A Deep Learning Algorithm for Automated Cardiac Murmur Detection Via a Digital Stethoscope Platform

**DOI:** 10.1101/2020.04.01.20050518

**Authors:** John S. Chorba, Avi M. Shapiro, Le Le, John Maidens, John Prince, Steve Pham, Mia M. Kanzawa, Daniel N. Barbosa, Brent E. White, Jason Paek, Sophie G. Fuller, Grant W. Stalker, Sara A. Bravo, Dina Jean, Subramaniam Venkatraman, Patrick M. McCarthy, James D. Thomas

**Affiliations:** Division of Cardiology, University of California San Francisco; Division of Cardiology, Zuckerberg San Francisco General Hospital; Eko, Oakland, CA; Division of Cardiology, Bluhm Cardiovascular Institute, Northwestern University

## Abstract

**Background:** There is variability among clinicians in their ability to detect murmurs during cardiac auscultation and identify the underlying pathology. Deep learning approaches have shown promise in medicine by transforming collected data into clinically significant information.

**Objective:** The objective of this research is to assess the performance of a deep learning algorithm to detect murmurs and clinically significant valvular heart disease using recordings from a commercial digital stethoscope platform.

**Methods:** Using over 34 hours of previously acquired and annotated heart sound recordings, we trained a deep neural network to detect murmurs. To test the algorithm, we enrolled 373 patients in a clinical study and collected recordings at the four primary auscultation locations. Ground truth was established using patient echocardiograms and annotations by three expert cardiologists.

**Results:** Algorithm performance for detecting murmurs has sensitivity and specificity of 76.3% and 91.4%, respectively. By omitting softer murmurs, those with grade 1, sensitivity increases to 90.0%. The algorithm detects moderate-to-severe or greater aortic stenosis with sensitivity of 97.5% and specificity of 77.7% and detects moderate-to-severe or greater mitral regurgitation with sensitivity of 64.0% and specificity of 90.5%.

**Conclusion:** The deep learning algorithm’s ability to detect murmurs and clinically significant aortic stenosis and mitral regurgitation is comparable to expert cardiologists. The research findings attest to the reliability and utility of such algorithms as front-line clinical support tools to aid clinicians in screening for cardiac murmurs caused by valvular heart disease.

## Introduction

The stethoscope is an iconic medical instrument nearly synonymous with Western medicine. It is easy to handle, inexpensive, and its use is universally accepted—even expected—as part of a physical examination. Yet for the stethoscope to be a useful tool, it requires that the provider both captures and correctly interprets the diagnostic sounds of the patient. Though nearly all providers can perform the act of auscultation with minimal training, interpretation of the collected data is difficult, even for experienced and specialized providers. Inter-observer reliability for identification of a classic auscultatory finding – the “murmur” – is fair at best^1,2^, and the provider’s ability to identify the true pathology underlying that finding is even worse^3^. Moreover, these challenges are further exacerbated by a noisy and rushed environment, which is the norm in modern practice. Despite the paucity of data in this area, anecdotally, these conclusions ring true for a wide spectrum of medical providers. Since cardiac auscultation remains a cornerstone of the physical exam, diagnostic assistance of its interpretation could therefore be of great use.

Classical teaching of the cardiac auscultatory exam, and murmurs in particular, centers around the diagnosis of valvular heart disease. Worldwide, valvular heart disease (VHD) is a major cause of mortality and reduced quality of life for tens of millions of patients worldwide^4–8^. As life expectancies increase, so does the prevalence of VHD in the elderly. Annual VHD fatalities have increased 2.8% each year in the US since 1979 and are projected to double over the next 25 years^5,9^. Many forms of VHD manifest with a prolonged asymptomatic period, which can be dangerous if not identified. Patients with asymptomatic severe aortic stenosis (AS), a common form of VHD, who do not undergo immediate aortic valve replacement have been shown to have an annual rate of sudden death of 3-13%^10^.

Echocardiography remains the gold standard for diagnosis of VHD, given its minimal physical risk and excellent test characteristics^11^. Yet echocardiography requires both highly trained sonographers to acquire the data, as well as interpretation of the images by trained cardiologists. Accordingly, echocardiography is expensive, with the total annual cost of $1.2B for Medicare enrollees alone^12^. Despite its widespread use, it also requires a pre-existing suspicion from the referring provider, and it may not be locally available for patients in medically underserved areas. Because VHD is associated with textbook auscultatory findings^13^, cardiac auscultation can be a widespread, fast, and inexpensive tool to improve access to VHD screening, facilitate earlier detection of VHD, and reduce the need for unnecessary echocardiography. To begin to address this problem, we investigated the utility of an electronic stethoscope platform to develop a deep learning algorithm to identify cardiac murmurs.

Deep learning approaches have shown impressive results towards problems in the medical field in recent years, utilizing imaging data such as radiologic studies^14^ and echocardiograms^15^ to develop interpretative algorithms. Interest in using deep learning for stethoscope sound analysis has also expanded in recent years, leading to applications in lung^16^ and heart^17^ sound classification. Indeed, an independently developed algorithm, focused on the binary distinction between pathology and normal, has tested favorably in a pediatric cohort, generating confidence in this approach^18^. Deep learning has also shown promise in utilizing auxiliary data unintended to be part of the original data set^19^, illustrating its ability to turn seemingly trivial data into useful information. Based on the traditional training of cardiologists to identify and triage valvular pathology from auscultatory characteristics, we hypothesized that a deep learning approach could be developed to perform similarly, if not better, than these specialty providers.

## Methods

### Algorithm Development

Eko’s heart murmur detection algorithm has been approved for FDA 510(k) clearance^20^ and is integrated with the Eko digital stethoscope and ECG software platform to assess single heart sound recordings. The algorithm was trained on recordings from a HIPAA-compliant collection of 400,000 audio recordings from Eko CORE and DUO electronic stethoscopes. The training set consisted of 5,878 de-identified audio recordings totaling over 34 hours from 5,318 unique patients. Recordings were initially randomly selected from the first 60,000 recordings collected and then sub-selected to ensure sufficient murmur examples to train the model. The training data quality is thus representative of what we expect in actual clinical use. The test set recordings used to measure classification performance of the trained algorithm are entirely separate and collected specifically for this purpose.

To complete the training set for a supervised learning problem, audio recordings and phonocardiograms (PCGs) were reviewed and labeled by one physician as one of three classes: no heart murmur, heart murmur, or poor signal. Recordings of lung sounds, noise, human speech, etc. were examples of data labelled poor signal.

The neural network model used for PCG classification uses a ResNet^21^ deep convolutional neural network architecture. Prior to being sent to the model, input recordings are filtered using an 8^th^ order Butterworth high-pass filter at 30 Hertz and downsampled to 2000 Hertz. The final output of the network is a three component probability distribution used to classify the recording.

The end-to-end algorithm makes a sequence of binary decisions to produce one of three possible outputs. First, it determines whether the recording is of sufficient signal quality to classify as murmur/no murmur, using the output from the neural network corresponding to ‘Poor Signal’ as a measure of signal quality. If the signal quality is found to be below a pre-specified threshold, then the recording is classified as ‘Poor Signal’. Otherwise, the classifier then determines whether the signal shows the presence of a ‘Heart Murmur’ or can be classified as ‘No Heart Murmur’ based on another set threshold. All model parameters and thresholds are fixed at training time.

To validate the end-to-end algorithm, separate expert clinicians were used to annotate a test set of 1774 recordings collected from 373 patients through an ongoing multi-site study. For the binary Good Signal / Poor Signal classification step, the algorithm output was compared to the annotations of signal quality qualitatively. For the final murmur detection step, algorithm output is compared to annotations of systolic and diastolic murmur presence using the primary analysis measures of sensitivity and specificity. As secondary analyses, sensitivity and specificity were calculated for PCG stratified by recording position and device, while sensitivity was calculated for PCGs with murmurs stratified by grade.

To show that the algorithm is detecting murmurs associated with clinically significant VHD, such as aortic stenosis and mitral regurgitation, we additionally compare predictions for a subset of the test set patients to echocardiographic assessment of VHD.

### Clinical Study Design

To obtain the data for testing, we undertook a cross-sectional study of subjects presenting to the echocardiography laboratory and structural heart disease clinics at the Northwestern Memorial Hospital (Chicago, IL) and UCSF Medical Center (San Francisco, CA) to obtain paired electronic stethoscope recordings with clinical echocardiography results. When possible, we pre-screened potential subjects for those with suspected valvular disease to enrich for our target population of cardiac murmurs. The protocol was approved as a minimal risk study by each of the institutional review boards of the participating sites, and all patients gave written informed consent.

### Stethoscope Recordings

Recordings of the phonocardiogram (PCG) were performed by trained study personnel for each subject in a standardized manner. Each subject underwent 15 second recordings at the four standard auscultation positions (aortic: 2^nd^ intercostal space, right sternal border; pulmonic: 2^nd^ intercostal space, left sternal border; tricuspid: 5^th^ intercostal space, left sternal border; mitral: 5^th^ intercostal space, mid-clavicular line). These recordings were obtained with the standard, clinically available Eko mobile application wirelessly-connected first to the Eko CORE Stethoscope, then to the Eko DUO. Recorded PCG and ECG data were saved as 16-bit, 4000 and 500 Hz-sampled WAV files, respectively, and were synced in real-time to a HIPAA-compliant cloud storage location and sent to the algorithms for analysis. Auscultatory recordings were reviewed by the study investigators for quality control.

### Phonocardiogram Annotations

Using a custom-made web platform, expert annotators listened to heart sound recordings with headphones while viewing a plot of the PCG. Expert annotators were cardiologists having completed fellowship training in cardiology and each with at least 10 years of clinical practice. Two sensory modalities helped annotators assess signal quality (1-5 scale with defined rubric) and systolic and diastolic murmur presence (true or false for each) and grade (Levine scales 1-6 or 1-4^22^). Since murmur grade was determined by recording only, systolic murmur grades 5 and 6 were not used. To establish a single set of ground truth labels for a recording we aggregated the responses of the three cardiologists. For a binary presence of each murmur type, we used a majority vote. For signal quality and murmur grade, both of which were optional, we used the median of the responses if there were three and the lessor if there were only two.

### Echocardiographic Data

Clinical echocardiogram reports followed American Society for Echocardiography (ASE) guidelines which allowed grading of VHD as follows: none, mild, moderate, or severe.^23,24^ The echocardiography laboratories at our study sites included additional categories of trivial, trivial-to-mild, mild-to-moderate, moderate-to-severe, and critical. The final clinical reports of the subject echocardiograms were de-identified and associated with study subjects at the study sites, with particular attention to the presence and degree of valvular stenosis and regurgitation of each of the four valves. For our disease screening evaluation, we define “clinically important” or “significant” VHD cases as those graded moderate-to-severe or worse, for this would encompass all levels of disease which could require timely intervention beyond serial monitoring. We define controls as subjects free of valvular, structural, or congenital heart disease, with no valvular regurgitation or stenosis beyond trivial or physiologic severity.

### Statistical Analysis

Data analysis and visualization was performed in Python using the standard packages numpy, pandas, seaborn, and matplotlib. Confidence intervals were computed by bootstrap rather than approximations which require assumptions about data distributions. To compare proportions, such as sensitivity, on different data samples, the ‘N-1’ chi-squared test was used for statistical significance.

## Results

### Validation Study Population

Of the 373 subjects, 362 had sufficient echocardiographic information for inclusion in the final analysis and their characteristics are summarized in the Table 1. The patient population tended towards an elderly population, was predominantly white, and nearly equally split in gender, consistent with cases of valvular heart disease seen at academic medical centers.

**Table 1:**
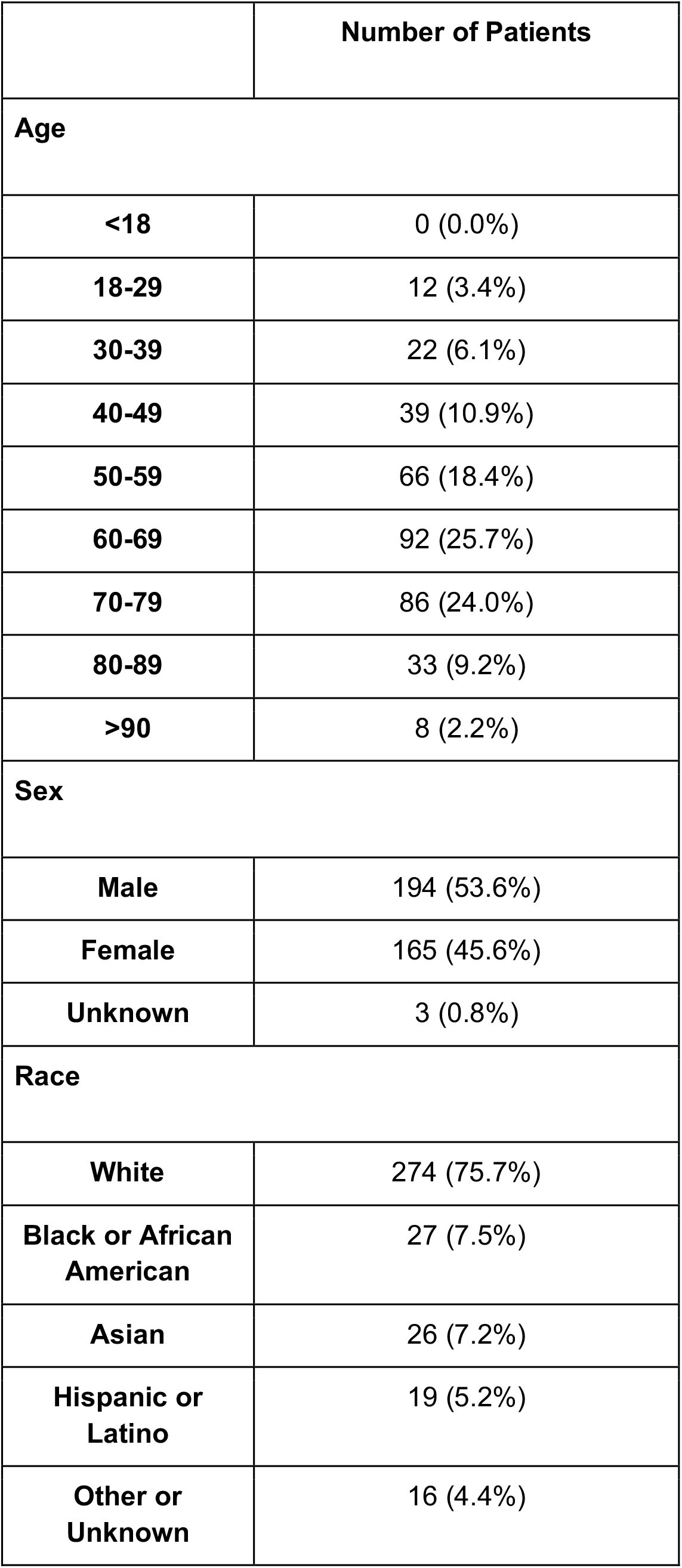
Characteristics of Study Patients

### Murmur Detection Performance

We first compared algorithm output on the test set of 1774 recordings to their annotated ground truth. The cardiologists provided a murmur/no murmur vote and ordinal grade to each of those recordings while the algorithm responded murmur/no murmur for only those of sufficient signal quality. To assess performance of this ‘poor signal’ screening, which excluded 226 recordings from subsequent murmur detection, we plotted median annotator sound quality histograms faceted by algorithm output in Figure 1. We observed that the ‘poor signal’ recordings have low annotator signal quality scores, showing the algorithm is not preventing analysis of potential murmurs when recordings are good. The algorithm was able to screen 1548 recordings or 87% of the test set, and thus would provide a useful clinical tool. All further performance analysis was based on this subset of the test set.

**Figure 1:**
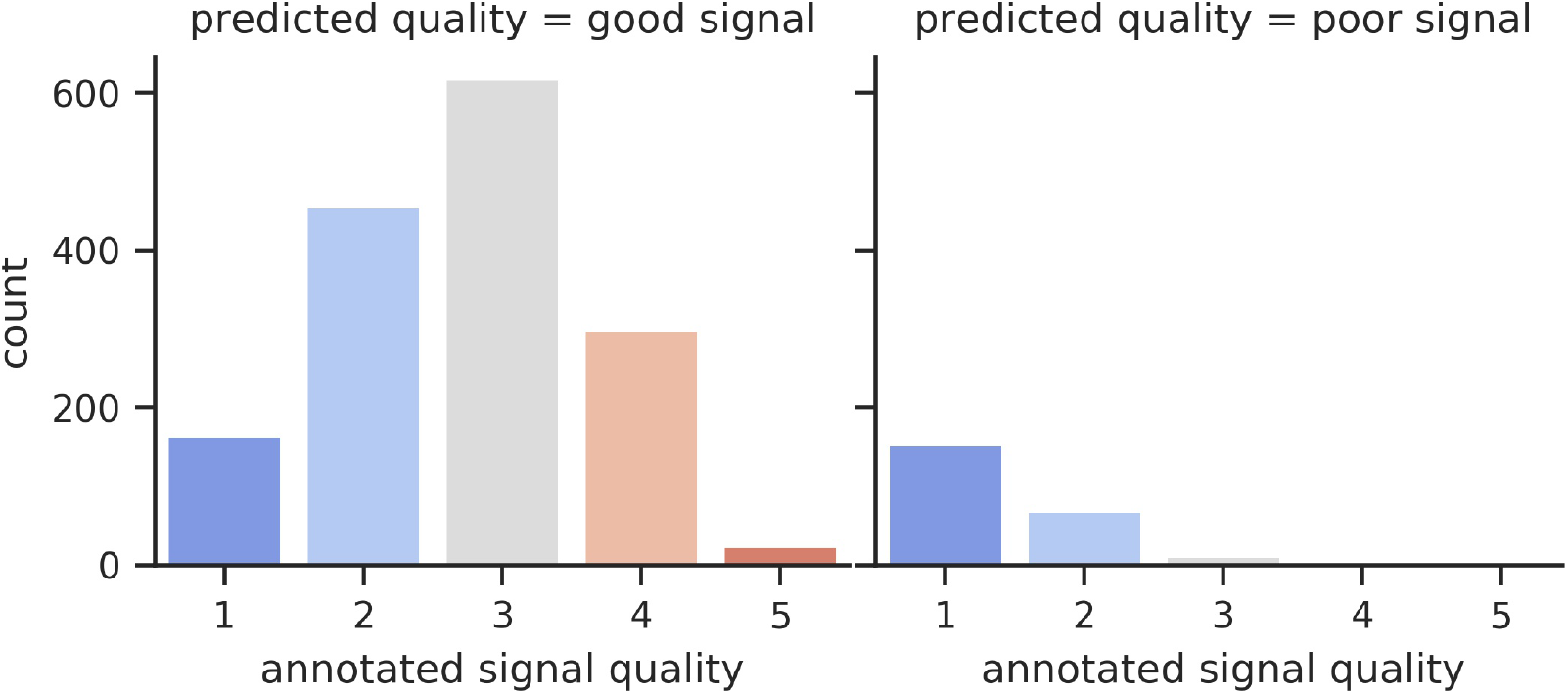
Predicted and annotated signal quality show favorable agreement. The plot on the right shows that the recordings predicted as ‘poor signal’ by the algorithm have low signal quality as assessed by the cardiologist annotators.

Filtering only the good signal recordings, we directly compared prediction to clinically defined ground truth in the confusion matrix in Table 2. Algorithm performance had a sensitivity and specificity for detecting murmurs of 76.3% (95% CI: 72.9–79.3%) and 91.4% (95% CI: 89.6– 93.1%), respectively (Table 3), and a positive predictive value of 86.6% (95% CI: 84.0–89.3%) using the murmur prevalence from this test set. The likelihood ratios positive and negative are 8.89 (95% CI: 7.35–11.08) and 0.259 (95% CI: 0.225–0.297), respectively (Table 3).

**Table 2:** Confusion matrix for single recording murmur detection using annotator ground truth.

|  | **No murmur detected** | **Murmur detected** | ***Total*** |
| --- | --- | --- | --- |
| **No murmur annotated** | 817 | 77 | 894 |
| **Murmur annotated** | 155 | 499 | 654 |
| ***Total*** | 972 | 576 | 1548 |

**Table 3:** Murmur detection sensitivity by cardiologist-assessed intensity grade

| **Murmur Grade** | **Number of recordings** | **Sensitivity (95% CI)** | **Specificity (95% CI)** | **Likelihood Ratio Positive (95% CI)** | **Likelihood Ratio Negative (95% CI)** |
| --- | --- | --- | --- | --- | --- |
| all | 654 | 76.3 (72.9–79.3) | 91.4 (89.6–93.1) | 8.86 (7.35–11.08) | 0.259 (0.225–0.297) |
| ≥2 | 451 | 90.0 (87.1–92.6) | 91.4 (89.6–93.0) | 10.5 (8.6–12.9) | 0.109 (0.081–0.141) |

Our algorithm sensitivity compared to annotations may appear low at least in part because the expert annotators were very sensitive themselves, able to hear very low grade murmurs. Omitting soft murmurs (annotator-aggregated grade 1), which are less likely to indicate meaningful disease^2,25,26^, sensitivity significantly increased to 90.0% (*p* < 0.0001, Table 3).

Next, we evaluated whether the performance of our algorithm differed based on auscultatory location. This analysis is summarized in Table 4. Overall, these performances were similar, as evidenced by the overlapping confidence intervals. Notably, recordings at the pulmonic position were more numerous in this subset since fewer recordings at that position were removed by the algorithm as ‘poor signal’.

**Table 4:** Algorithm performance by auscultation position

| **Position** | **Number of Recordings** | **Sensitivity** | **Specificity** | **Likelihood Ratio Positive** | **Likelihood Ratio Negative** |
| --- | --- | --- | --- | --- | --- |
| aortic | 382 | 75.0 (68.5–81.4) | 89.4 (84.6–93.4) | 7.1 (4.9–11.4) | 0.28 (0.21–0.35) |
| mitral | 388 | 71.2 (63.7–78.4) | 92.7 (89.4–96.0) | 9.7 (6.7–17.7) | 0.31 (0.24–0.39) |
| pulmonic | 450 | 81.9 (76.6–86.9) | 91.1 (87.3–94.6) | 9.2 (6.4–14.7) | 0.2 (0.14–0.26) |
| tricuspid | 328 | 75.4 (68.0–82.4) | 92.4 (88.7–95.7) | 10.0 (6.6–17.8) | 0.27 (0.19–0.35) |

The recordings are made from two devices, the Eko CORE Stethoscope and the Eko DUO, which also records a 1-lead ECG. We thus evaluated whether the performance of the algorithm differed based on the specific device used to capture the sounds. Detection performance for each device is given in Table 5, with sensitivity on CORE recordings slightly but significantly higher than on those from DUO (*p* < 0.05). However, when controlling for signal quality (Table 6), the algorithm is not consistently more sensitive on CORE recordings, an example of the well-known Simpson’s Paradox.

**Table 5:** Algorithm performance by recording device

| **Device** | **Number of Recordings** | **Sensitivity** | **Specificity** | **Likelihood Ratio Positive** | **Likelihood Ratio Negative** |
| --- | --- | --- | --- | --- | --- |
| CORE | 929 | 77.6 (73.7–81.4) | 91.2 (88.5–93.6) | 8.8 (6.7–12.1) | 0.25 (0.2–0.29) |
| DUO | 619 | 73.2 (65.9–79.7) | 91.6 (88.7–94.1) | 8.7 (6.4–12.3) | 0.29 (0.22–0.37) |

**Table 6:** Algorithm performance by signal quality and recording device. CORE and DUO sensitivities are only significantly different at *p* < 0.05 for recordings with signal quality equal to 2.

| Signal Quality | Device | Number of Recordings | Sensitivity |
| --- | --- | --- | --- |
| 1 | CORE | 35 | 50.0 (0.0–100.0) |
|  | DUO | 128 | 66.7 (0.0–100.0) |
| 2 | CORE | 231 | 63.3 (48.5–76.9) |
|  | DUO | 227 | 52.6 (35.7–68.6) |
| 3 | CORE | 408 | 70.9 (65.5–77.1) |
|  | DUO | 203 | 76.1 (68.4–84.4) |
| 4 | CORE | 238 | 89.5 (84.4–93.8) |
|  | DUO | 56 | 82.9 (69.4–94.1) |
| 5 | CORE | 17 | 100.0 (100.0–100.0) |
|  | DUO | 5 | 100.0 (100.0–100.0) |

Allowing the algorithm’s positive-negative decision boundary to vary, we can plot a receiver operating characteristic (ROC) curve to show different sensitivity and specificity tradeoffs. The FDA-cleared murmur detection algorithm, however, operates at a single point on this ROC curve, with performance described above. Figure 2 shows this ROC curve with the operating point of Eko software overlaid. Stratification of the ROC curve based on grade of the murmur again shows that the detection algorithm operates with significantly improved characteristics with a higher grade murmur (Fig. 2, green line).

**Figure 2:**
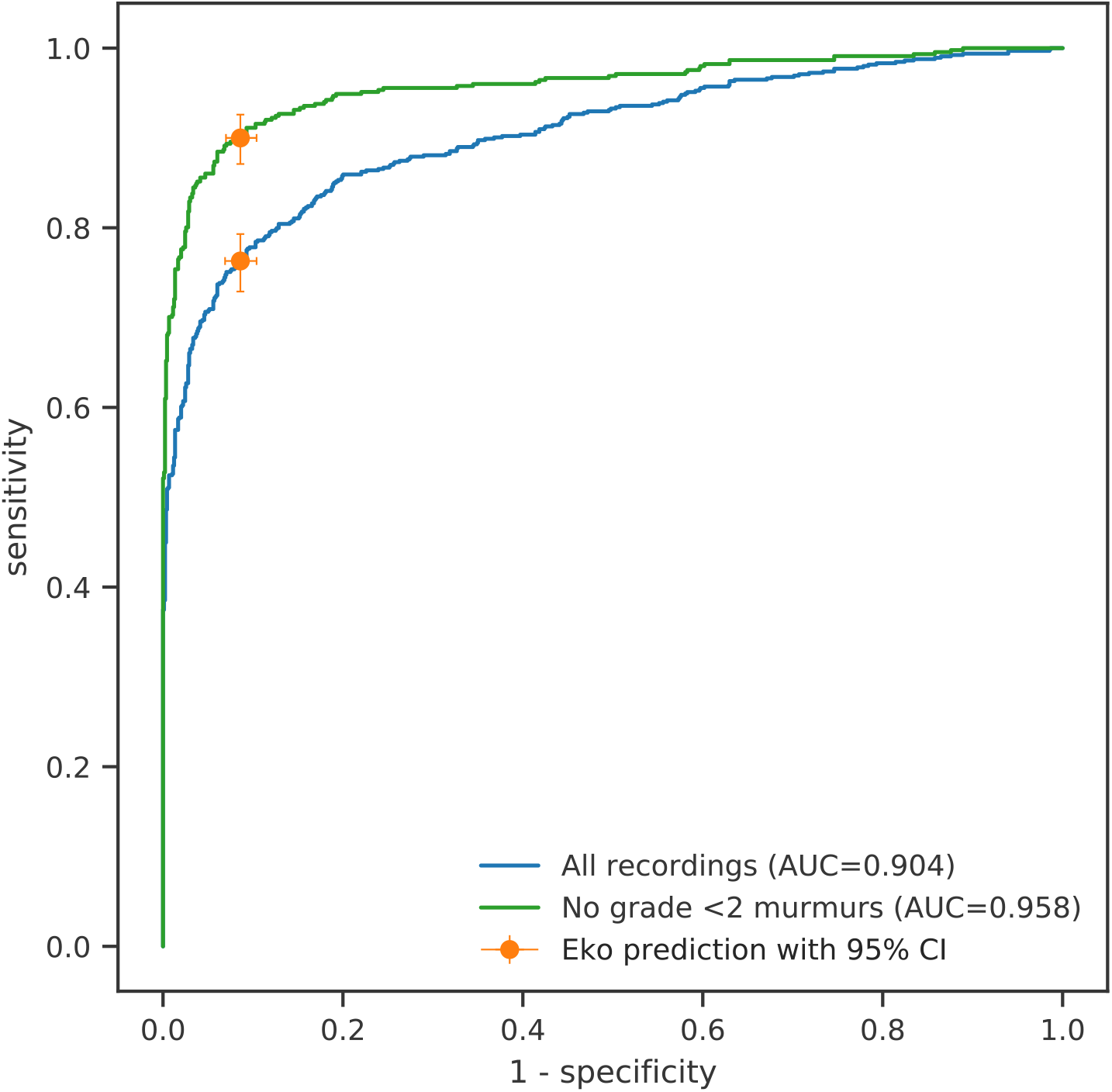
Performance of murmur detection algorithm. ROC curves for all recordings (blue) and minimal intensity-filtered murmurs (green) are show. Eko software operates with parameters yielding the orange marker. Error bars indicate 95% confidence intervals.

### Valvular Disease Screening Performance

Having evaluated the algorithm on individual recordings, we can measure its performance as a patient screening tool for valvular heart disease by comparing predictions to echocardiograms. First, we consider aortic stenosis (AS). From the 373 patients represented in the test set, we exclude 60 patients with valve replacements and 11 with insufficient echocardiogram reports. Of the remaining, we group 40 having aortic stenosis of severity at least moderate-to-severe as cases and 103 patients without structural heart disease (*i.e*. no valve disease greater than trivial severity, and no congenital disease), as controls. As mentioned previously, the severity threshold for disease was chosen to include cases which would require evaluation for mechanical intervention. Figure 3 illustrates the flow of study participants. Importantly, whereas for the murmur detection algorithm the “gold standard” is simply the clinical interpretation (and likely more subject to inter-observer variability), for valvular heart disease screening, the “gold standard” is the echocardiogram. Since an aortic stenosis murmur is generally loud, we defined a positive test as one where a murmur is detected at either the aortic or pulmonic positions. A negative test is one where no murmur is detected at the aortic and pulmonic positions. A small fraction of subjects in both patient groups (i.e. cases and controls) had recordings limited by a ‘poor signal’ classification and were removed from the analysis if necessary.

**Figure 3:**
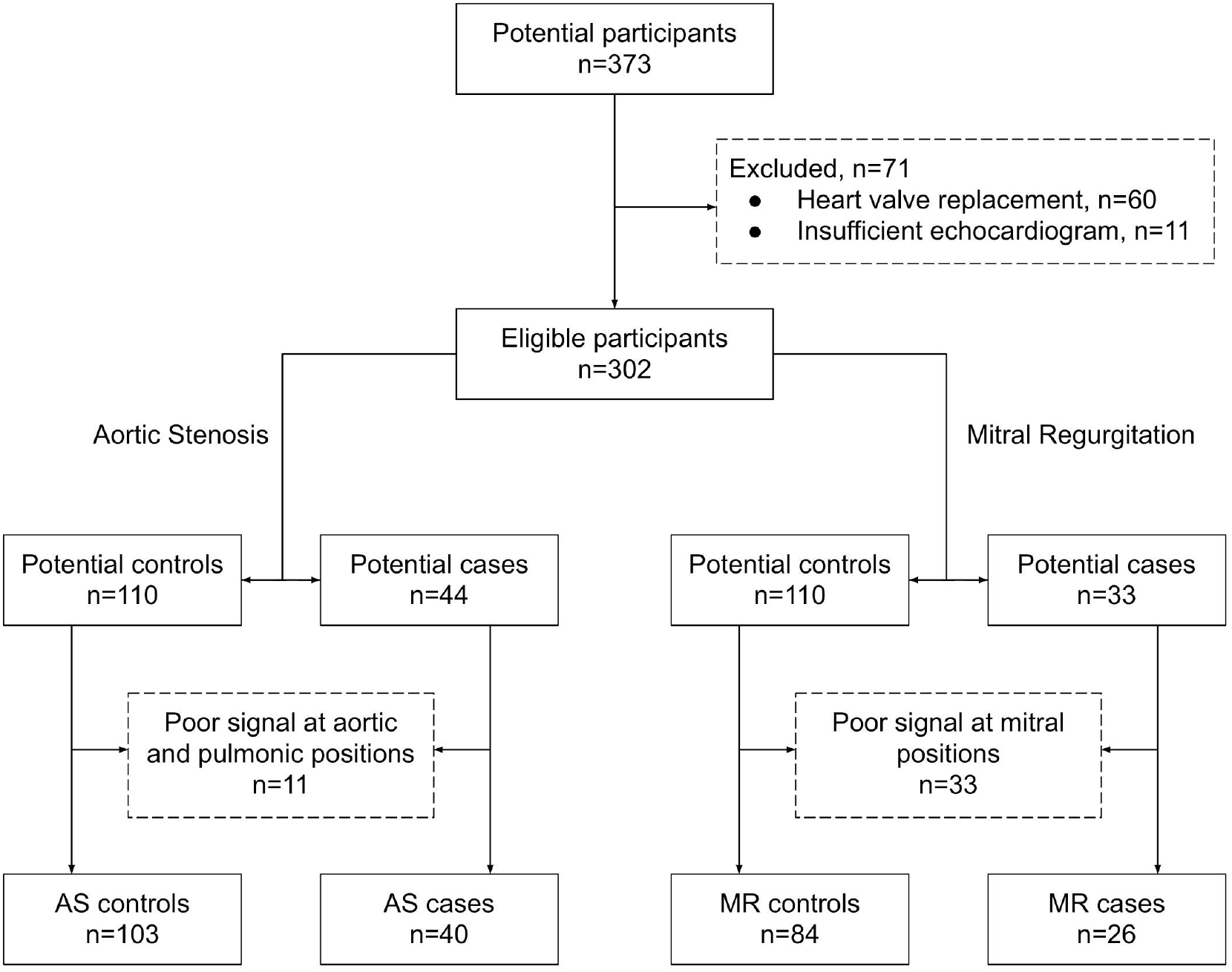
Flow of VHD study participants. We defined VHD cases as those graded moderate-to-severe or worse to encompass all levels of disease which could require timely intervention beyond serial monitoring. We defined controls as subjects free of valvular, structural, or congenital heart disease, with no valvular regurgitation beyond trivial or physiologic severity.

We show an ROC curve of the test results for aortic stenosis detection in the 143 subjects in Figure 4. The single points plotted are taken from Table 7. Overall, the Eko murmur detection algorithm in production compares favorably to the expert clinicians, with higher sensitivity than all annotators and higher specificity than two of the three.

**Table 7:** Aortic Stenosis screening results on 40 cases and 103 controls.

|  | **Sensitivity**<br>**(CI)** | **Specificity**<br>**(CI)** | **Likelihood Ratio**<br>**Positive (CI)** | **Likelihood Ratio**<br>**Negative (CI)** |
| --- | --- | --- | --- | --- |
| **Algorithm** | 97.5 (91.7–<br>100.0) | 77.7 (69.2–<br>85.3) | 4.37 (3.16–6.63) | 0.032 (0.0–0.108) |
| **Annotator A** | 97.5 (91.7–<br>100.0) | 42.7 (33.0–<br>52.0) | 1.7 (1.44–2.05) | 0.059 (0.0–0.2) |
| **Annotator B** | 90.0 (80.0–<br>97.8) | 77.7 (69.1–<br>85.3) | 4.03 (2.86–6.16) | 0.129 (0.029–0.261) |
| **Annotator C** | 87.5 (76.5–<br>97.3) | 91.3 (85.4–<br>96.3) | 10.0 (5.9–24.2) | 0.137 (0.03–0.255) |

**Figure 4:**
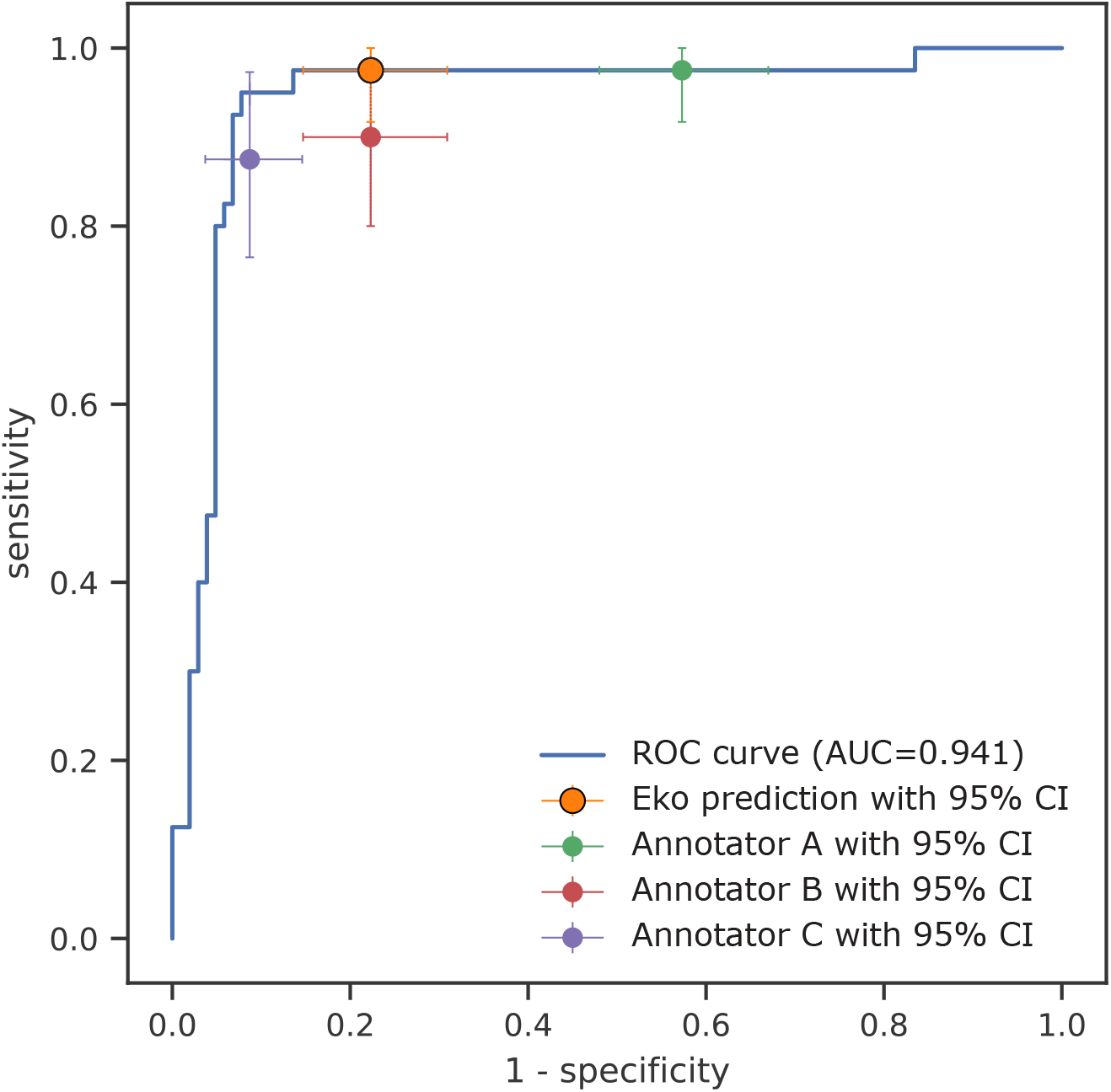
Performance of aortic stenosis screening by murmur detection algorithm. The algorithm ROC curve is shown in blue. Eko software operates at the orange marker. The performance of the individual cardiologists is shown by the green, red, and purple markers. Error bars indicate 95% confidence intervals.

We are also able to screen for mitral regurgitation (MR) with the murmur detection algorithm. Using the same patient cohort, and the same inclusion and exclusion criteria as for aortic stenosis, except testing at the single mitral/apex location, we have 32 cases and 84 controls. There are fewer control subjects for MR because there are a greater number of ‘poor signal’ recordings at the mitral position. The ROC curve is presented in Figure 5. The algorithm compares favorably to the annotators, whose performances fall along the ROC curve. Performance metrics with confidence intervals are given in Table 8.

**Table 8:** Mitral regurgitation screening results on 32 cases and 84 controls,

|  | **Sensitivity**<br>**(CI)** | **Specificity**<br>**(CI)** | **Likelihood Ratio**<br>**Positive (CI)** | **Likelihood Ratio**<br>**Negative (CI)** |
| --- | --- | --- | --- | --- |
| **Algorithm** | 65.4 (46.5–83.3) | 90.5 (83.6–96.4) | 6.87 (3.58–18.9) | 0.383 (0.185–0.595) |
| **Annotator A** | 84.6 (69.6–96.7) | 63.1 (52.5–73.6) | 2.29 (1.66–3.29) | 0.244 (0.05–0.499) |
| **Annotator B** | 69.2 (52.0–86.3) | 79.8 (71.3–87.8) | 3.42 (2.18–5.87) | 0.386 (0.169–0.614) |
| **Annotator C** | 65.4 (47.1–83.3) | 86.9 (79.3–93.7) | 4.99 (2.87–10.75) | 0.398 (0.192–0.618) |

**Figure 5:**
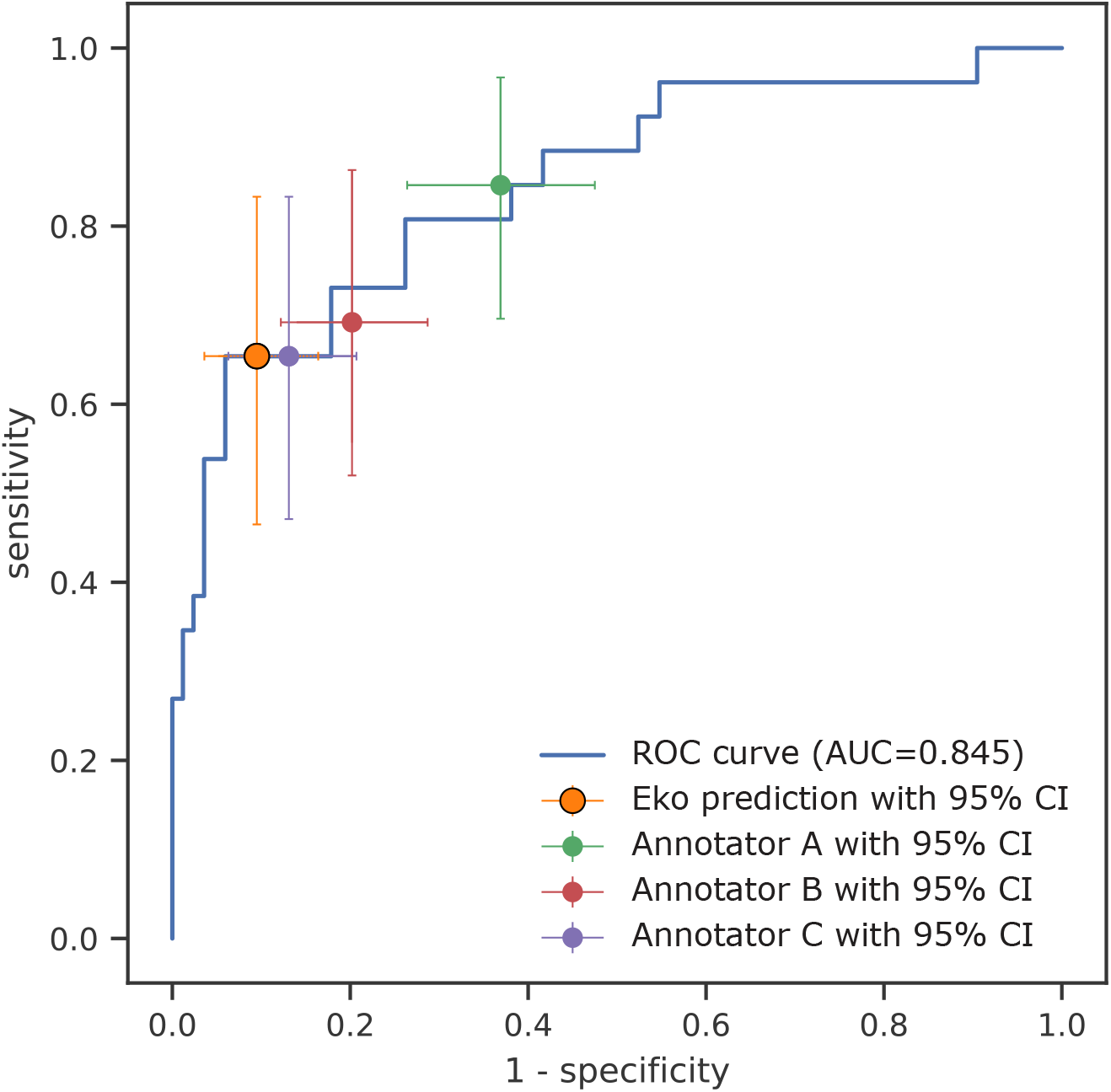
Performance of mitral regurgitation screening by murmur detection algorithm. The algorithm ROC curve is shown in blue. Eko software operates at the orange marker. The performance of the individual cardiologists is shown by the green, red, and purple markers. Error bars indicate 95% confidence intervals.

## Discussion

Our results show that within our test set, the algorithm detects a murmur of any grade with 76.3% sensitivity and 91.4% specificity and detects moderate-to-severe or greater aortic stenosis with a sensitivity of 97.5% and specificity of 77.7%. These numbers are more sensitive than all the expert cardiologist annotators and more specific than all but one of them. The algorithm detects moderate-to-severe or greater mitral regurgitation with a sensitivity of 64.0*%* and a specificity of 90.5%, which is more specific but less sensitive than the annotators. Notably, as “algorithms” themselves, the annotators fall nearly directly on the ROC curve of the deep learning algorithm, as applied to mitral regurgitation. As seen for both detection of aortic stenosis and mitral regurgitation, there is considerable variability in performance amongst annotators, even amongst highly trained experts. This likely represents variability in real-world practices amongst clinicians, which is a considerable challenge. This emphasizes the need for a real-time tool that could obviate this inter-observer variability, and the algorithm tested here, when paired with an electronic stethoscope, does exactly this.

The algorithm presented here is intended as a decision support tool for clinicians, and our data support its use in detecting murmurs attributed to valvular heart disease. To put this in perspective, in the general elderly population, where the prevalence of surgically intervenable aortic stenosis is approximately 5%^27^, a negative test, carrying a negative likelihood ratio of 0.032, will nearly rule out the diagnosis, reducing its probability to <0.2%. Conversely, applying the positive likelihood ratio of 4.37 in the setting of a positive result will increase the disease probability to 19%, though this would include non-surgical VHD disease given our study design. In both cases, however, these changes will almost assuredly affect clinical management. Moreover, it is likely that the overall accuracy to make a clinical diagnosis of VHD would be higher when combining the provider’s interpretation of the heart sounds along with the algorithm results^28^. We anticipate that such a tool would be particularly useful in a hurried setting like an emergency department, where minimizing the time to diagnostic test results, as well as the strain on providers, is particularly important. While an emergent environment was not explicitly captured in our test set, we purposefully captured heart sounds in a real-world clinical setting to enhance the generalizability of our findings.

Importantly, the algorithm tested here is designed solely for the evaluation of the presence of a cardiac murmur in single recordings. Auscultatory findings, however, are much richer than simply the presence or absence of a murmur. Indeed, classical teaching of aortic stenosis includes non-murmur characteristics, such as the “softness” of the A2 component of the second heart sound, and the timing of the peak of the systolic murmur, as indicators of severity^13^. It is reasonable to conjecture that an extended algorithm, which includes more known signs of VHD in its decision-making process would therefore have better performance as a complete disease screening test. This is an area of active investigation and development for our group as low inter-observer reliability in the identification of other murmur characteristics, or even patients with specific diseases, creates many challenges for curating richly labeled datasets large enough for training comprehensive heart sound models. This, however, underscores the need for more investigation in this area. Though the test set populations may well represent the U.S. population, they may not reflect populations in developing countries, where the prevalence and etiology of VHD are different. As these populations would likely very much benefit from a potentially low-cost support tool such as this, further investigation in these populations are warranted.

Our results also show that the cardiologists and the algorithm perform better in detecting aortic stenosis than detecting mitral regurgitation. This may be attributed to aortic stenosis having a more discernible auscultatory signature. Moreover, its characteristic murmur can be heard in both the aortic and pulmonic positions on auscultation, offering additional data points for both the human and computer decision-makers. Physiologically, mitral regurgitation is a load-dependent murmur, such that severity of regurgitation can vary markedly depending on the minute-to-minute hemodynamics of the patient. Moreover, MR can be directional, and therefore certain types of MR may simply not have murmurs at a single auscultation location, like the cardiac apex. The higher rates of poor signal from the mitral position in our study only exacerbate these challenges. Further study of additional auscultation locations, or alternative, non-murmur auscultatory characteristics, would be useful to aid in improving the performance of VHD detection algorithms.

The tested algorithm addresses the need for an effective, reliable, and accessible method to screen for murmurs and ultimately, detect VHD. First, the deep learning algorithm is accurate and reliable, with low inter-operator variability. It shows comparable performance to that of an expert cardiologist, suggesting it could deliver cardiology-level expertise to frontline clinicians in the field. Potential benefits include enabling clinicians to detect VHD earlier and more consistently, and potentially reduce morbidity and mortality due to earlier clinical intervention^29^. Second, the algorithm can detect heart murmurs at the point of care through cellular or wi-fi connectivity with the stethoscope and mobile platform. A tool that can serve as an affordable alternative for diagnosing VHD would be very valuable since echocardiography remains limited by cost, time, and access, especially in rural and underserved areas suffering from a shortage of cardiologists^30^. Third, in light of the recent and ongoing COVID-19 pandemic^31^, the potential to provide expert level diagnostics through telemedicine, thereby limiting the transmission of a highly contagious disease, is also particularly attractive. Furthermore, the digital stethoscope platform employed here could be extended to investigate other auscultation findings, such as lung sounds, which may help improve screening for such disease. Fourth, to the extent the algorithm can accurately exclude valvular disease, it could reduce the burden of unnecessary echocardiography. Overall, our study shows the promise of this tool as an adjunct to clinical care, and illustrates the potential of it expanding into something even greater.

The deep learning algorithm evaluated here is designed to be used as a clinician-assisted screening tool. Similar to blood test results, the algorithm can have false positive and false negative results, and therefore it is important that results of the algorithm are placed within the appropriate clinical context. We note that the murmur-detection algorithm was developed using, to our knowledge, the world’s largest adult echocardiogram-paired heart sound recording database. Looking ahead, this database can potentially be used in the development of other clinical tools, including algorithms to differentiate between innocent and pathologic murmurs, as well as algorithms to identify specific types of VHD, such as aortic stenosis, mitral regurgitation, and tricuspid regurgitation.

In summary, valvular heart disease has become an increasingly prevalent manifestation of poor cardiovascular health in both the developed and developing world. A clinical decision support algorithm that is able to detect and classify valvular heart disease could improve public health, reducing unnecessary referrals for echocardiography and promoting early and accurate screening in underserved areas. Using an electronic stethoscope platform, we have deployed the first stage of such a tool: a deep learning algorithm to identify cardiac murmurs as manifestations of valvular heart disease.

## Data Availability

Additional supporting data are available upon request from the corresponding authors. Programming code related to data processing and not subject to intellectual property or confidentiality obligations will be made available upon request. All requests for raw and analyzed and related materials data will be reviewed by the corresponding authors and the Eko legal department to verify whether the request is subject to intellectual property or confidentiality obligations. Any data and materials that can be shared will be released via a Material Transfer Agreement. Patient-related data not included in the paper were generated as part of an ongoing prospective clinical study (NCT03458806) and may be subject to patient confidentiality and IRB review.

## Acknowledgements

This project was supported by NIH R43HL144297 (to JM and JSC) and K08HL124068 (to JSC). JDT was supported in part by a grant from the Irene D. Pritzker Foundation.

## Author Contributions

Conception and design: JSC, JM. Data acquisition: DNB, MMK, BEW, JaP, SGF, GWS, SAB, DJ. Data analysis: AMS. Data interpretation: JSC, AMS, LL, JM, SP, SV, BEW, PMM, JDT. Software creation: AMS, JM, DNB, JoP, SV. Manuscript generation: AMS, LL, JSC. Manuscript revision: JSC, AMS, LL, JM, SP, SV, JDT.

